# Immunity in Omicron SARS-CoV-2 breakthrough COVID-19 in vaccinated adults

**DOI:** 10.1101/2022.01.13.22269213

**Authors:** Hassen Kared, Asia-Sophia Wolf, Amin Alirezaylavasani, Anthony Ravussin, Guri Solum, Trung The Tran, Fridtjof Lund-Johansen, John Torgils Vaage, Lise Sofie Nissen-Meyer, Unni C. Nygaard, Olav Hungnes, Anna H Robertson, Lisbeth Meyer Næss, Lill Trogstad, Per Magnus, Ludvig A Munthe, Siri Mjaaland

**Author notes:** These authors contributed equally to this work (Ludvig A Munthe, Siri Mjaaland). Corresponding authors: Hassen Kared, Ludvig A Munthe.

## Abstract

The new SARS-CoV-2 variant of concern (VOC) Omicron has more than 30 mutations in the receptor binding domain (RBD) of the Spike protein enabling viral escape from antibodies in vaccinated individuals and increased transmissibility^1-6^. It is unclear how vaccine immunity protects against Omicron infection. Here we show that vaccinated participants at a superspreader event had robust recall response of humoral and pre-existing cellular immunity induced by the vaccines, and an emergent *de novo* T cell response to non-Spike antigens. We compared cases from a Christmas party where 81 of 110 (74%) developed Omicron breakthrough COVID-19^7^, with Delta breakthrough cases and vaccinated non-infected controls. Omicron cases had significantly increased activated SARS-CoV-2 wild type Spike-specific (vaccine) cytotoxic T cells, activated follicular helper (T_FH_) cells, functional T cell responses, boosted humoral responses, activated anti-Spike plasmablasts and anti-RBD memory B cells compared to controls. Omicron cases had significantly increased *de novo* memory T cell responses to non-Spike viral antigens compared to Delta breakthrough cases demonstrating development of broad immunity. The rapid release of Spike and RBD-specific IgG^+^ B cell plasmablasts and memory B cells into circulation suggested affinity maturation of antibodies and that concerted T and B cell immunity may provide durable broad immunity.

---

On Nov 25, 2021, a new SARS-CoV-2 variant of concern (VOC), Omicron (B.1.1.529), was reported by South Africa^1,2^. Omicron has more than 30 mutations in the Spike protein and 15 in the receptor binding domain (RBD) of the Spike protein that binds to the uptake receptor ACE2. These include shared mutations with the Alpha, Beta, Gamma, and Delta VOCs^2,5,6,8^. These previously well-studied mutations confer increased transmissibility, higher viral binding affinity, and higher antibody escape^3,4^. Other mutations are novel and of unknown significance, but reports suggest increased transmission in South Africa^2^, and a 40-fold reduced effect of IgG neutralizing antibodies in Pfizer BNT162b2–vaccinated patients as analyzed in Omicron VOC neutralization assays^9^. The Omicron VOC has since been found in over 89 countries as of mid-January 2021.

In contrast to the serological epitopes, bioinformatic analyses suggest that most immunodominant T cell epitopes are conserved^10^. We aimed to test if cases had fully functional vaccine generated T cell responses and to provide in-depth analyses of the immune responses in Omicron breakthrough COVID-19.

We found that the Omicron and Delta breakthrough COVID-19 cases were all mild and did not require hospitalization. 6-13 days after symptoms debut, no major changes were found in lymphocyte, monocyte or granulocyte counts (data not shown). The cases had near normalized acute phase proteins and systemic inflammatory markers (Extended data Figure 1 and data not shown). However, the Omicron infected had a signature of increased CXCL4 (PF4, released during platelet activation), MPO, GDF-15 and Galectin-9 (linked to tissue damage), and sCD14 and LBP (associated with monocyte activation), Extended data Figure 1. We hypothesized that the resolution of inflammation was due to the pre-existing adaptive immunity induced by the vaccine.

To determine the activation of vaccine T cell immunity, we detected CD8^+^ T cells specific for Spike peptides with MHC-class I restricted multimers (See Methods). We also performed deep immune-phenotyping to detect markers that correlate with protective immune responses in mild COVID-19, and quantified cells (see below). High dimensionality analyses of Omicron cases revealed that nearly all Spike-specific CD8^+^ T cells were activated expressing HLA-DR, CD38^11^, CD27 and PD-1^12^ (Figure 1a-b and Extended Data Figure 2). In contrast, Delta breakthrough cases had lower frequency of activated cells, non-activated cells had markers of terminal differentiated effector functions (KLRG1, GPR56)^13,14^, Figure 1a-b and Extended Data Figure 2. Thus, clustering showed expression of activation/memory markers in Omicron COVID-19, while Delta showed increased levels of effector markers^13-15^,.

**Figure 1.**
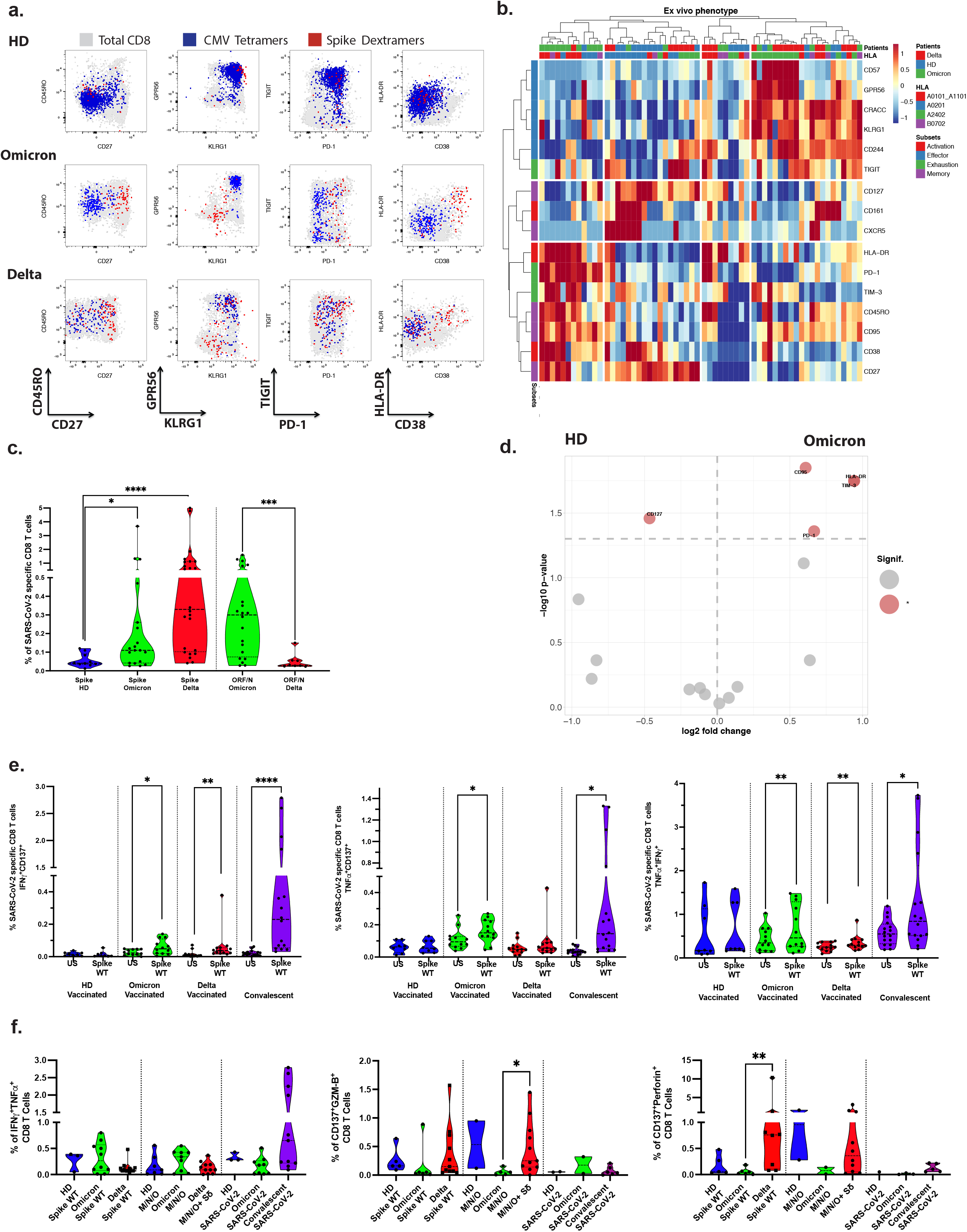
Cytotoxic cellular immunity during Omicron and Delta breakthrough infection. a. Characterization of Spike (S)-, and CMV-specific CD8^+^ T cells. Anti-Spike Dextramer (red) and anti-CMV tetramer staining (blue) were combined with deep immune-phenotyping by flow cytometry and overlaid on total CD8 T cells (grey). See Methods for peptides and peptide: HLA-multimers. b. *Ex vivo* immune phenotype of SARS-CoV-2 Spike-specific CD8^+^ T cells. A cold to hot heatmap represents the scaled frequency of each individual marker expressed by antigen-specific CD8^+^ T cells. The distribution of markers and patients was automatically performed by unsupervised hierarchical clustering. Patient, HLA type and marker subsets are indicated in the top two rows and in the leftmost column respectively. c. Quantification of SARS-CoV-2 Spike and non-Spike specific CD8^+^ T cells defined by peptide: HLA-multimers. d. Signature of Spike-specific CD8 T cells. The specific markers are based on the fold change and significance on the volcano plot. e. Functionality of Spike specific CD8^+^ T cells. PBMCs were stimulated overnight *in vitro* by overlapping SARS-CoV-2 Spike (wildtype, WT) peptides and stained for activation markers, dual expression (over background) of IFN-g, CD137 and TNF is shown. f. Responses in vitro to Spike (WT) peptides, left; middle: pooled overlapping SARS-CoV-2 membrane (M) and nucleoprotein (N) peptides, and ORF peptides (See Methods Table S2) – with additional mutated VOC Delta Spike (Sd) peptides for the Delta patients, and right: the whole proteome of WT SARS-CoV-2 (S, M, N, E, O), see Methods. Cytotoxicity markers are shown. See also Extended data Figure 2&5a.

Similarly, principal component analysis biplots showed that activation markers such as CD95, HLA-DR, PD-1^12^, and negative feedback inhibitory TIM-3 characterized the Omicron COVID-19 response, while Delta responses showed influence of terminal effector molecules such as CD57 and CD244^16^. Both had significant reduction of CD127, a marker found in the steady state (Figure 1d, Extended Data Figure 2).

The phenotype and frequency of T cells directed against cytomegalovirus (CMV) or Epstein-Barr virus (EBV) were unaltered during Omicron and Delta breakthrough COVID-19 (Figure 1a and Extended Data Figure 2), therefore excluding systemic bystander activation. In summary, the Delta breakthrough COVID-19 profile was reminiscent of vaccine boost responses, while the Omicron profile resembled the more potent T cell activation found in infected, unvaccinated individuals.

Quantification of Spike-specific CD8^+^ T cells in Omicron- and Delta-infected cases showed a significantly increased frequency as compared to vaccinated healthy controls (p= 0.0248, and p<0.0001 respectively) (Figure 1c), but a lower frequency in Omicron than in Delta breakthrough COVID-19 (p= 0.0349).

We next turned to the development of *de novo* peptide: HLA multimer binding CD8^+^ T cells to the rest of the virus (i.e. non-Spike proteins). We found a marked expansion and activation towards non-Spike peptides (ORF3a, ORF1ab and Nucleocapsid, see Methods). Moreover, Omicron cases had increased frequency of non-Spike specific CD8^+^ T cells compared to Delta cases (p=0.0004), and these had a memory phenotype, contrasting with an effector phenotype found in Delta cases (Figure 1d and Extended Data Figure 2).

As a corollary to the expanded and activated state of vaccine-specific CD8^+^ T cells, we found that the Omicron breakthrough cases had significantly increased frequency of *in vitro* CD8^+^ T cell responses to Spike peptides, including significantly increased CD137^+^ cells that secreted IL-2, IFN-γ, and TNF (Fig 1e). Functional responses from SARS-CoV-2 specific CD8^+^ T cells were barely detectable in vaccinated healthy donors (3-4 months post-vaccine) but were still strong in convalescent patients (3-6 months post recovery).

We next analyzed functional CD8^+^ T cell responses to other (non-Spike) antigens of the SARS-CoV-2 virus. Omicron and Delta breakthrough COVID-19 cases had Membrane-, Nucleoprotein- and ORF-specific T cells but lower responses than observed in recovered convalescent patients (Figure 1f). Hence, strong vaccine-specific CD8^+^ T cell immunity was accompanied by emergent immunity to SARS-CoV-2 epitopes that included a multitude of epitopes. CD8^+^ T cells from Delta-infected patients also presented increased cytotoxic properties as assessed by the expression of Granzyme-B or perforin (Figure 1f). The augmented functionality may be explained by the terminal effector profile of these Spike SARS-CoV-2 specific CD8 T cells (Figure 1a-b, Extended Data Figure 2).

Among the other T cell subsets, we observed a peripheral decrease of TCRγ/δ^+^ T cells, and no significant modulation of MAIT cells or CD4/CD8 ratio (data not shown). In the T helper (Th) subsets, we did not observe any general upregulation of markers associated with T cell differentiation, activation, exhaustion or senescence. However, we next evaluated whether T follicular helper (T_FH_) cells were modulated, a subset that is involved in humoral responses (in germinal center for B cells maturation) but also can have cytotoxic properties in COVID-19 hospitalized patients^17^. An unbiased mass cytometry analysis identified T_FH_ cellular clusters in representative donors (Figure 2a and Extended Data Figure 3a-b). These included T_FH_ clusters increased in Omicron (Figure 2b) expressing PD-1 (important for B cell help, clusters C2, C3), proinflammatory marker such as CD161 (C7, C16) or activation markers such as CD38 or HLA-DR (C9), see also Extended Data Figure 3. Finally, we extended our high dimensional analysis to the entire cohort for the markers of interest and performed supervised analysis. Mass cytometry study (confirmed by flow cytometry staining, data not shown) revealed the decreased frequency of T_FH_ cells in the Omicron-infected cases (p=0.0071) combined with an activated profile (up-modulation of CD95 and HLA-DR, p=0.0456 and p=0.0119 respectively but with the down-regulation of chemokine receptors associated to Th1 (CXCR3), Th2 or Th17 response (CCR4 and CCR6), p= 0,0387, p= 0.0124 and p= 0.0096 respectively, Figure 2c). The impact of this T_FH_ profile on isotype switching and the affinity maturation of B cells remains to be investigated. It is possible that ongoing T cell-B cell collaboration and retention of T_FH_ cells in lymph nodes could explain this decrease in T_FH_ frequency. Nevertheless, the functionality of vaccine-induced Th cells was enhanced in Omicron cases in comparison to vaccinated controls (Figure 2d). The acquisition of activation-induced markers (CD154, CD137) and the production of cytokines was stronger in Omicron-infected cases (p=0.0485 and p=0.0075 for CD137 and IFN-γ respectively). The ability to secrete TNF (p=0.0199) and IL-2 (and indirectly to proliferate, (p=0.0372) was enhanced in Omicron versus Delta breakthrough patients. However, a detectable response against non-spike (combined with the Delta VOC mutated Spike) was observed only in the Delta patients.

**Figure 2.**
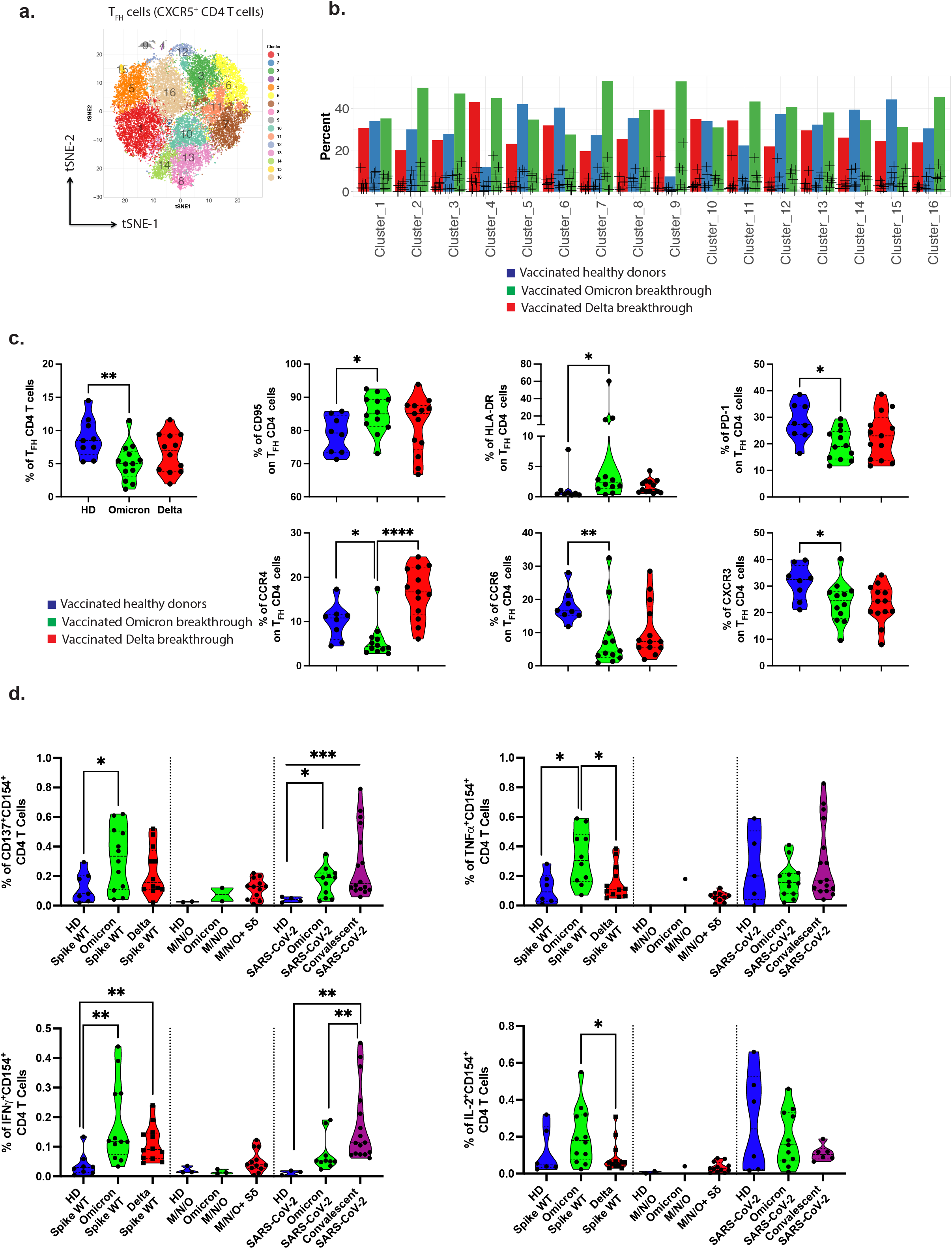
Helper cellular immunity during Omicron and Delta breakthrough infection. a. Mass Cytometry profile of Follicular helper CD4 T cells. Visualization by tSNE-plots of clusters automatically identified by Phenograph in total T_FH_ from concatenated files of vaccinated HD, Omicron-infected, and Delta-infected (2000 cells from 6 individuals per group). b. Cluster distribution of T_FH_ CD4 cells. Relative proportion of each cluster is represented for each group of patients. See Extended Data Figure 3a-c for markers in clusters. c. Characterization of T_FH_ CD4 cells. The frequency of T_FH_ was quantified in cryo-preserved PBMCs by mass cytometry and represented for all patients by violin plots. d. Functionality of SARS-CoV-2 specific CD4^+^ T cells. The specific responding CD4^+^ T cells are shown similarly as in Fig.1f. See also Extended data Figure 5b.

In contrast to the Delta breakthrough COVID-19 cases (p=0.001), the frequencies of anti-Spike B cells and anti-RBD spike B cells were not increased in Omicron COVID-19 breakthrough cases. However, IgG anti-RBD was significantly increased in Omicron suggesting a boost of previous vaccine responses (p=0.0061), but less so than in Delta breakthrough (p=0.0023) (Figure 3a-b and Extended Data Figure 4). Furthermore, Delta cases had a relatively increased responsiveness to RBD in comparison to healthy donors or Omicron cases (p=0.0213 and p=0.0117 respectively). Isotype switched Spike binding B cells (not expressing IgM or IgD) were significantly more likely to express IgG in Omicron than in Delta cases. This applied both for anti-RBD and anti-Spike (non-RBD), with p=0.0062 and p=0.0215 respectively, (Extended Data Figure 4). As expected, the concentration of anti-RBD IgG correlated significantly with the frequency of Spike-specific B cells (p=0.0003), increased significantly with time post-infection (p=0.0004), and the age of the participants was negatively associated with the humoral response in our small cohort (p=0.0189), Extended Data Figure 4.

**Figure 3.**
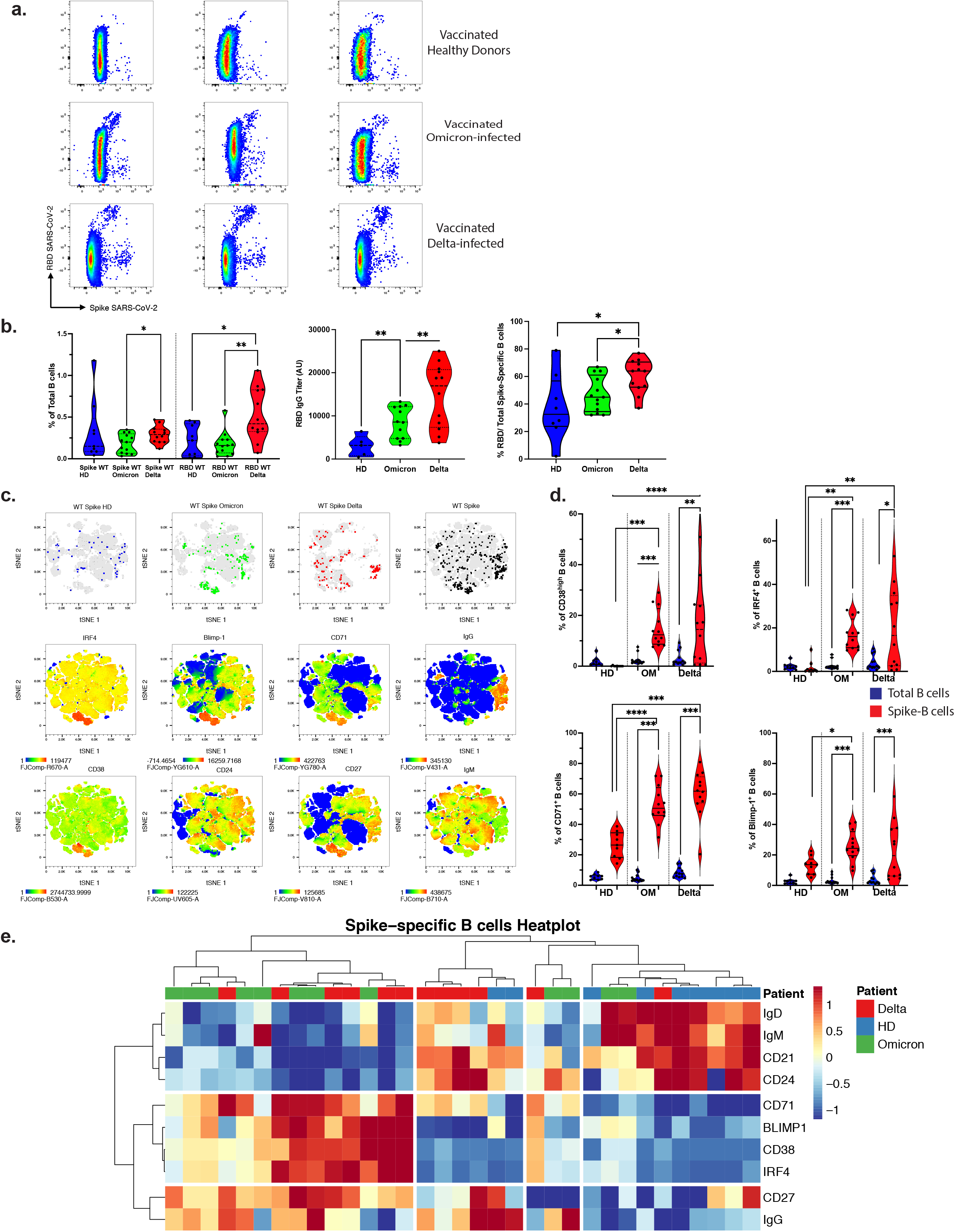
Humoral and B cell immunity during Omicron and Delta breakthrough infection. a. Identification of Spike-binding and doubly Spike- and RBD-binding B cells in total B cells from three cases: Healthy Donor (HD), Omicron breakthrough and Delta breakthrough. b. Quantification of Spike-binding and RBD-binding B cells. Left: Absolute frequency of Spike-binding and RBD-binding B cells. Middle: serum levels of anti-RBD IgG (Arbitrary Units/ml). Right: Relative frequency of anti-RBD to spike specific B cells. Ratio of RBD-binding/total Spike-binding (RBD+ non-RBD Spike) B cells is shown. c. Phenotype of Spike-binding B cells. Visualization by tSNE-plot of selected markers as indicated (bottom two rows) in total B cells and location of Spike-binding B cells (top row) in HD, Omicron, Delta and aggregated donors (20 000 cells from two individuals per group, total=120 000 cells). Each marker is visualized by a cold to hot heatmap. d. Quantification of anti-Spike antibody secreting cells (ASC). The frequency of specific markers for ASC among B cells (CD38, IRF4, CD71, BLIMP-1) are shown for Total B cells (blue) and Spike-binding B cells. e. Characterization of anti-Spike B cells. A heat-plot shows the phenotype of Spike-specific B cells in the three groups. The normalized frequency of each marker is displayed and automatic hierarchical clustering of Spike-binding B cells for each patient is shown. See also Extended data Figure 4&5c

The most striking B cell response in Omicron COVID-19 cases was significantly increased Spike-specific plasma blasts (CD38^hi^CD27^hi^CD21^low^CD24^neg^CD71^hi^ IRF4^+^BLIMP-1^+^) where about half were IgG^+^ (Figure 3 c-e and data not shown). The Spike-specific B cells also included activated IgG^+^CD24^+^CD21^+^ memory cells (CD27^+^CD71^+^, Figure 3e, Extended Data Fig. 4). The data demonstrate a surge of vaccine-specific differentiated plasmablasts and activated memory B cells that were similarly developed in the Delta and Omicron breakthrough COVID-19

We here describe robust T cell activation towards SARS-CoV-2 vaccine peptide antigens in breakthrough infections of cases with the SARS-CoV-2 Omicron VOC. The T cell phenotype corresponded to successful protective T cell immunity in COVID-19 convalescents including activation markers and lack of exhaustion markers. A more potent activation was seen in Omicron than in the Delta breakthrough cases where T cell phenotypes were consolidated (in terms of effector markers) rather than activated. As a corollary, *in vitro* challenge with SARS-CoV-2 Spike peptides demonstrated a significantly increased T cell responsiveness for both CD4^+^ T helper cells and CD8^+^ T cells. We also found a robust emergent T cell immunity towards other SARS-CoV-2 non-vaccine peptides suggesting development of broad T cell responses. This was accompanied by a significantly boosted IgG anti-RBD response and a surge of IgG^+^ RBD and Spike specific B cells that were plasma cell precursors or activated memory B cells.

In the current study, as many as 74% of the participants developed Omicron COVID despite being fully vaccinated. Preliminary analyses^9^ suggest that the increased transmission and breakthrough infections relate to escape of antibody neutralization as sera from Pfizer BNT162b2–vaccinated individuals had dramatically reduced viral neutralization. In addition, neutralizing antibodies (NAbs) peak and decay with time after vaccination.^18^ Most infected individuals at the Christmas party were young to mid-aged adults that had been vaccinated 3-6 months before the party, suggesting some decay of NAb titers.

If Omicron escapes protective antibodies, exposed individuals will rely on the collaboration between vaccine expanded T cell and B cell responses to counteract the Omicron infection. Bioinformatic analyses suggest that Omicron mutations do not impact immunodominant Spike peptide antigens and that protective vaccine-generated T cell responses may be triggered by the Omicron infection^10^. The current results confirm this perspective.

Importantly, in contrast to decaying antibody titres, SARS CoV-1 T cell memory was long-lasting and was found after 17 years.^19^ The importance of T cells has been underlined in previous reports as T cells are necessary for rapid and efficient resolution of COVID-19, for protection against severe COVID-19 in settings of low antibody levels, and for rapid viral control in the absence of antibodies - aborting infection in healthy individuals.

The current results demonstrate a potent activation of vaccine-specific T cell responses in Omicron breakthrough COVID-19 that contrast with consolidated effector markers seen in the responses in Delta breakthrough infection. It is possible that the relative lack of neutralizing antibodies provides a greater antigenic challenge and activation of T cells in the Omicron cases, thereby resembling the more potent T cell activation found in unvaccinated individuals. The re-challenge may have implications for durability of the T cell responses.

mRNA-based vaccination induces a robust SARS-CoV-2-specific antibody response, and a SARS-CoV-2-specific germinal centre B cell and T_FH_ cell response^20,21^. Moreover, vaccination generates responses that included functional Th cells and class switched memory B cells that cross-bind Alpha, Beta, and Delta RBDs, and are capable of rapidly producing functional antibodies after stimulation^22^. The current results demonstrate rapid generation of Spike/RBD-binding plasmablasts and activated IgG^+^ memory cells in the Omicron breakthrough COVID-19 as a result of prior vaccination. It is likely but remains to be demonstrated that the B cell responses will provide adapted anti-Omicron NAbs as a result of the germinal centre response and that concerted T and B cell immunity as seen here will provide broad and long-term protection against SARS-CoV-2.

## Data Availability

All data produced in the present study are available upon reasonable request to the authors

## Acknowledgments

We would like to thank Ingvild Bokn and Kari Mette Midtdal for participation in data collection, Ingrid Egner, Katrine Persgård Lund, Viktoriia Chaban, Julie Røkke Osen, Marit Fodnes Killengreen, Åse-Karine Fjeldheim, Tove Karin Herstad, Linn Margrethe Eggesbø, Thea Kristine Rogne Møller and Ingrid Fadum Kjønstad for biobanking and technical help. The Dep of Medical Biochemistry at the Oslo University Hospital is thanked for analysing acute phase reactants. The Dept of Microbiology at the Oslo University Hospital is thanked for providing PCR cycle threshold (Ct) values from diagnostic SARS-CoV2 PCR samples.

## Author contributions

HK, LAM and SM conceived and designed the study. HK and LAM drafted the paper. ASW, AR and SM contributed to drafting the paper. HK, ASW, AA, GS, TTT performed experiments; HK, ASW, AA, GS, TTT, LAM, and SM analysed and interpreted data, AR, GS, JTV, LSNM, UCN, OH, AHR, LMN, LT, PM contributed to data analysis and interpretation. SM, LSNM, JTV, AHR, LMN, LT, PM organized collection of samples and information from cases. All authors contributed to and approved the final manuscript.

## Supplementary Information

is available for this paper.

Correspondence and requests for materials should be addressed to Hassen Kared, Ludvig A Munthe

## Methods

### Cases and controls

We included four main cohorts: 1) participants from the Christmas party with Omicron VOC COVID-19 (n=13, mild disease) as described^1,^ 2) donors with breakthrough Delta VOC COVID-19 (n=13, mild disease) from the Norwegian Institute of Public Health (NIPH) Young Adult cohort (see Table 1 and Extended Data Fig. 1 for overview of cases), 3) control fully vaccinated health care workers (healthy donors, HD) at the Oslo University Hospital (n=14), and 4) samples from SARS-CoV-2 (Wuhan-Hu-1)-infected COVID-19 convalescent blood bank donors who were bio-banked 3-6 months post-recovery in June-August 2020 and served as positive controls (n=16). Donors signed informed consent forms. Ethical approvals have been obtained (Approval numbers: REK 2021.233704; REK 2020.135924; REK 2021.229359).

The cases with the Omicron breakthrough COVID-19 were collected as part of the routine outbreak investigation performed by the Norwegian Institute of Public Health (NIPH). Subjects with Delta variant of concern (VOC) breakthrough COVID-19 were recruited from the Young Adult Cohort, NIPH. This cohort was established at NIPH and designed to understand the consequences of the pandemic among subjects aged 18-30 years. Samples were sequenced to confirm infection by the Omicron or Delta VOC (as designated by the World Health Organization). One of the original 14 Delta VOC breakthrough COVID-19 controls was disqualified as a control since sequencing revealed infection by Omicron and not Delta VOC. This sample was transferred to the Omicron cases for a total of n=13.

**Supplementary Table 1.** Overview of COVID-19 Cases.

|  | **Delta**<br>**breakthrough**<br>**COVID-19**<br>**Controls (n=13)** | **Omicron**<br>**breakthrough**<br>**COVID-19 Cases**<br>**(n=13)\*** | **Noninfected, HD**<br>**health care workers**<br>**(n=14)** |
| --- | --- | --- | --- |
| **Age (y)** |  |  |  |
| mean (range) | 27 (21-30) | 37.3 (28-50) | 31.9 (25-44) |
| median | 27 | 38 | 31 |
| **Sex** |  |  |  |
| Female, n (%) | 8 (61.5) | 9 (69.2) | 10 (71.4) |
| Male, n (%) | 5 (38.5) | 4 (30.8) | 4 (28.6) |
| **Vaccine type, dose 2** |  |  |  |
| BNT162b2<br>(Pfizer-BioNTech), n<br>(%) | 8 (61.5) | 8 (61.5) | 14 |
| mRNA-1273<br>(Moderna), n (%) | 5 (38.5) | 5 (38.5) | 0 |
| **Time since infection (d)** |  |  |  |
| mean (range) | 12.3 (3-22) | 9.9 (6-13) | - |
| median | 11 | 10 | - |
| **Time since dose 2 vaccination (d)** |  |  |  |
| mean (range) | 147 (92-297) | 117 (94-171) | 115 (92-167) |
| median (range) | 115 | 112 | 115 |
| **Time between dose 1 and 2 (d)** |  |  |  |
| mean (range) | 49 (27-83) | 46 (36-66) | 68 (64-87) |
| median | 46 | 43 | 64 |
| **Vaccine type, dose 1** |  |  |  |
| BNT162b2<br>(Pfizer-BioNTech), n<br>(%) | 9 (69.2) | 8 (61.5) | 14 |
| mRNA-1273<br>(Moderna) n (%) | 4 (28.6) | 4 (30.8) | 0 |
| **Previous COVID-19** |  |  |  |
| n (%) | 0 | 1 (7.7) | 0 |

\* one of the Young Adult cohort was found to be infected with the Omicron variant and was subsequently included with the other Omicron cases.

All participants had received both doses of SARS-CoV-2 vaccines (either BNT162 or mRNA-1273) according to the Norwegian National Vaccination Program. The two vaccines were given with an interval of 4-12 weeks (median of 6.5 weeks). Subjects had received their second vaccine doses between 13 and 42 weeks prior to this outbreak (with a median of 16 weeks and mean of 21 weeks).

Information on vaccination and infection were obtained from the Norwegian Immunization Registry (SYSVAK) and Norwegian Surveillance System for Communicable Diseases (MSIS).

### Sample preparation and HLA typing

All participants were screening for HLA typing using the freshly isolated PMBC samples. Cells were stained with anti-HLA-A2 (Biolegend), HLA-A24 (LSBio) and HLA-B07(Biolegend) and typed by flow cytometry. Individuals matched positive for these HLA were subsequently stained with the corresponding Dextramers/ Tetramers Class I restricted (HLA-A*02:01, HLA-A*24:02, and HLA-B*07:02 respectively). Negative patients for one of the HLA-A screened were directly tested for HLA-A*01:01, HLA-A*11:01 by Dextramer staining directed against SARS-CoV-2 and bystander virus (CMV, EBV, Influenza, see overview of peptides below in section on specific memory CD8 T cells). Sample within the groups had similar HLA distribution with HLA A02 and A24 in >5/13 cases, and B17 in more than 3 cases.

Each sample consisted of 2 aliquots with an average cell number of 10 million cells and a viability above 95% per donor. Samples were thawed at 37°C and immediately transferred into complete RPMI medium (10% FCS, 1% penicillin /streptomycin, glutamine, 10 mM HEPES). After the first wash, Thawed cells were incubated during 15 minutes at room temperature with DNAse (STEMCELL). Live cells were purified by removing dead cells using a column-based magnetic depletion approach according to the manufacturer’s recommendations (Miltenyi). Vaccinated healthy donor PBMCs matched for at least one of the donor HLA alleles were included in each experiment as control for specific T cell identification. VeriCells were included in each experiment as control for phenotypic markers.

### Flow cytometry

Thawed peripheral blood mononuclear cells (PBMCs) were stained with antibody panels to quantify and phenotype specific T cell responses to Spike peptides and B cell responses to RBD or Spike protein. Cells were acquired on a BD FACSymphony (BD Biosciences) or Attune NxT (ThermoFisher).

The following mAbs and stains were utilized for acquisition on BD FACSymphony: BB515 Mouse Anti-Human CD279 (PD-1) Clone EH12.1, BD Biosciences, PerCP-eFluor 710, KLRG1 Monoclonal Antibody (13F12F2), eBioscience, PE/Cyanine7 anti-human GPR56, Clone CG4, Nordic Biosite, Alexa Fluor 700 anti-human CD244 (2B4), clone C1.7, Nordic Biosite, APC/Cyanine7 anti-human HLA-DR, clone L243, Nordic Biosite, BV480 Rat Anti-Human CXCR5 (CD185) (Clone: RF8B2) BD Biosciences, BB515 Mouse Anti-Human CD38, clone, HIT2 BD Biosciences, Brilliant Violet 570™ anti-human CD3, Nordic Biosite, Brilliant Violet 605, CD127 Mouse anti Human, Clone HIL 7R M21, BD Biosciences, Brilliant Violet 650, CD161 Mouse anti Human, clone: DX12, BD Biosciences, BV711 Mouse Anti-Human TIM-3 (CD366), clone 7D3, BD Biosciences, BV750 Mouse Anti-Human CD8, clone SK1, BD Biosciences, Brilliant Violet 785™ anti-human CD57 Recombinant, clone QA17A04, Nordic Biosite, BV421 Mouse Anti-Human CD319 (CRACC), BD Biosciences, BUV395 Mouse Anti-Human TIGIT, clone 741182, BD Biosciences, Live/dead™ Fixable Blue Dead Cell Stain Kit, for UV excitation, Thermo Fisher Scientific, BUV563 Mouse Anti-Human CD45RO, clone UCHL1, BD Biosciences, BUV615 Mouse Anti-Human CD95, clone DX, BD Biosciences, BUV661 Mouse Anti-Human CD4, clone SK3, BD Biosciences, BUV737 Mouse Anti-Human CD38, clone HB7, BD Biosciences, BUV805 Mouse Anti-Human CD27, clone L128, BD Biosciences. VeriCells PBMC (BioLegend) were included as controls.

Frequency values were calculated based on the percentage of the parent immune cell population and phenotypic markers were gated individually for each sample and calculated as % of positive cells. High-dimensional phenotypic profiles and sample distributions were shown using uniform manifold approximation and projection. Data analysis was performed using CYTOGRAPHER® (ImmunoScape cloud based analytical software), custom R-scripts, GraphPad Prism (GraphPad Software) and FlowJo v10 software (BD Life Sciences). Statistical significance was set at a threshold of *p < 0.05, **p < 0.01, and ***p < 0.001.

### In vitro stimulation assays

Thawed cells were stimulated for 16h with SARS-CoV-2 PepTivator Spike protein peptides (Wuhan-Hu-1, Miltenyi Biotec). For non-Spike (WT) responses cells were stimulated with Nucleoprotein (PepTivator SARS-CoV-2 Prot N) and Membrane protein (PepTivator SARS-CoV-2 Prot M), consisting of 15-mer sequences with 11 amino acid overlaps in addition to the 4 ORF1ab/Orf3a peptides in Table S2. i.e. stimulated with M+N+O, Figure 1f (middle). The Delta breakthrough samples were in addition stimulated with mutated Spike peptides (PepTivator SARS-CoV-2 Prot S AY.1), i.e stimulated with M+N+O and + mutated Delta Spike. Alternatively, in Fig 1f (right), cells were stimulated with 88 pooled immunodominant oligopeptides from the whole proteome (PepTivator SARS-CoV-2 Select, Miltenyi Biotec) consisting of peptides from structural proteins (S, M, N, E) as well as non-structural proteins (O). Peptide stimulation was performed in the presence of costimulatory antibodies against CD28 and CD49d (BD Biosciences) and Brefeldin-A (10 μg/mL, Millipore Sigma). SARS-CoV-2-specific T cells were identified by dual expression of CD40L (CD154) and CD137, interferon-gamma (IFN-γ), interleukin-2 (IL-2), or tumour necrosis factor (TNF) for CD4^+^ T cells and by dual expression of IFN-γ and TNF or CD137 and IL-2, TNF or IFN-γ for CD8^+^ T cells.

### Detection of specific memory CD8 T cells

Antigen-specific CD8 T cells were detected by peptide: HLA multimers (see Supplementary Table S2 for overview). Spike Specific CTL were detected using PE-conjugated Dextramers targeting Spike and restricted to HLA-A*0101 (LTDEMIAQY^2-7^), HLA-A*0201 (YLQPRTFLL^3,5,8-11^), HLA-A*2402 (QYIKWPWYI^3-5,7,8^), and HLA-B*0702 (SPRRARSVA^5,9,12^). The panel was expanded using Flex-T tetramer according to the manufacturer’s instructions. We UV-exchanged peptides for Spike epitopes restricted to HLA-A*0101 (YTNSFTRGVY^2,5^), HLA-A*0201 (LITGRLQSL^11-14^ and RLNEVAKNL^6,11,12,15^), HLA-A*2402 (NYNYLYRLF^2,3,7,16^), and HLA-B*0702 (APHGVVFL^3,6^) and tetramerized with Streptavidin-PE (Biolegend). Similar approach was performed for non-Spike derived epitopes, including HLA-A*0101 (ORF3a, FTSDYYQLY^3,5,12,17^ and ORF1ab, TTDPSFLGRY^4-6,8^), HLA-A*0201 (ORF3a, LLYDANYFL^3,8,9,12^), HLA-A*2402 (ORF3a, VYFLQSINF^3,4,8^), and HLA-B*0702 (Nucleoprotein, SPRWYFYYL^8,9^) and tetramerized with Streptavidin-APC (Biolegend). CMV-and EBV/FLU-specific CD8 T cells were generated similarly and tetramerized using Streptavidin-PECF594 (Biolegend) and Streptavidin-PE-Cy5 respectively. CMV derived epitopes were for HLA-A*0101 (DNA polymerase processivity factor, VTEHDTLLY^3,7,18^), HLA-A*0201 (65 kDa phosphoprotein, NLVPMVATV^3,19^), HLA-A*2402 (65 kDa phosphoprotein, QYDPVAALF^3,20^), and HLA-B*0702 (65 kDa phosphoprotein, RPHERNGFTVL^3,21^) and EBV derived epitopes were for HLA-A*0201 (EBV LMP2, FLYALALLL) and HLA-B*0702 (EBV antigen 3, RPPIFIRRL^3,22^). A Flu peptide was for HLA-A*0101 (Nucleoprotein, Influenza A virus CTELKLSDY^23^). All peptides were ordered from Genscript with a purity above 85% by HPLC purification and mass spectrometry. Lyophilized peptides were reconstituted at a stock concentration of 10 mM in DMSO.

Antigen-specific multimer CD8 T cells were identified by fine manual gating.The designation of bona fide antigen-specific T cells was further dependent on (a) the detection cut-off threshold (≥ 5 events to be detected), (b) the background noise (frequencies of specific CD8^+^ T cells must be greater than frequencies from the corresponding CD4^+^ T cell population) as unbiased objective criteria for antigen-specificity assessment. Spike and non-Spike Dextramers staining have been extensively validated in COVID-19 convalescent patients and in SARS-CoV-2 vaccinated healthy donors during longitudinal follow-up.

**Supplementary Table 2.**
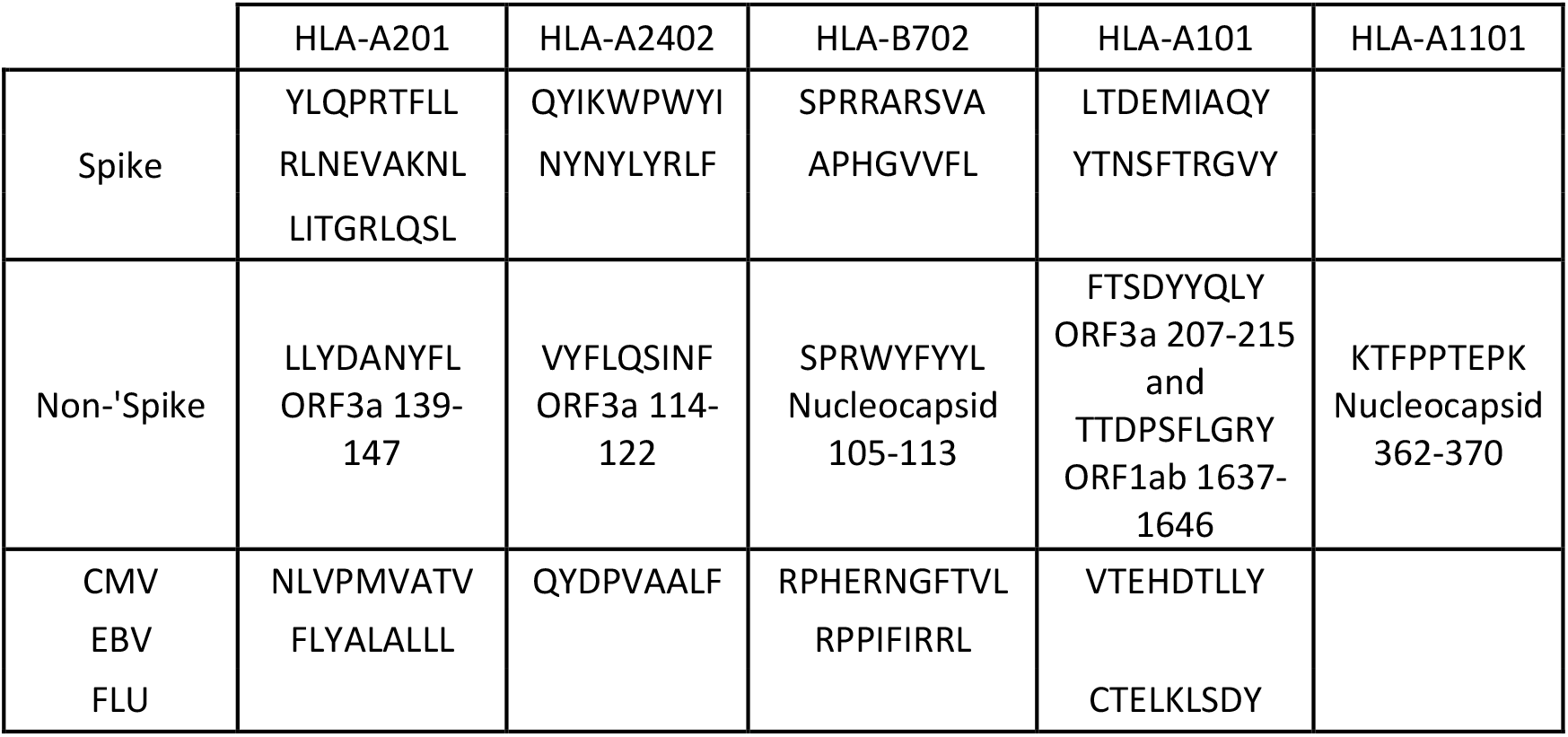
Peptide: HLA multimers.

### Mass Cytometry

#### Antibody staining panel setup

Purified CD19 (Biolegend), TCRVα7.2, CD160 and KLRG1 (R&D Systems) antibodies lacking carrier proteins (100 μg/antibody) were conjugated to DN3 MAXPAR chelating polymers loaded with heavy metal isotopes following the recommended labelling procedure (Fluidigm). The rest of the following mAbs were directly purchased from Fluidigm: y89 anti-CD45, 106Cd anti-CD45, 110Cd anti-CD45, 111Cd anti-CD19, 112Cd anti-CD45, 114Cd anti-CD45, 116Cd anti-CD45, 141Pr anti-CCR6, 142Nd anti-CD57, 143Nd anti-CD45RA, 144Nd anti-CD38, 145Nd anti-CD4, 146Nd anti-CD8, 147Sm anti-CD20, 148Nd anti-CD14, 149Sm anti-CD25, 150Nd anti-TCRVα7.2, 151Eu anti-lambda, 152Sm anti-TCRγδ, 153Eu anti-TIM-3, 154Sm anti-CD3, 155Gd anti-CD27, 156Gd anti-CXCR3, 158Gd100 anti-CCR4, 159Tb anti-TIGIT, 160Gd anti-Kappa, 161Dy anti-CD160, 162Dy anti-CD95, 163Dy anti-CRTH2, 164Dy anti-CD161, 165Ho anti-CD127, 166Er anti-CD85j, 167Er anti-CCR7, 168Er anti-CD71, 169Tm anti-NKG2A, 170Er anti-HLA-DR, 171Yb anti-CXCR5, 172Yb anti-KLRG1, 173Yb anti-CD141, 174Yb anti-PD anti-1/CD279, 176Yb anti-CD56, 209Bi anti-CD16. All labelled antibodies were titrated and tested by assessing relative marker expression intensities on relevant immune cell subsets in commercial lyophilized PBMCs from healthy donors (VeriCells, BioLegend). Antibody mixtures were prepared freshly and filtered using a 0.1 mM filter (Millipore) before staining.

#### Sample staining and acquisition

Cryo-preserved PBMCs were enriched for live cells by magnetic depletion of dead cells (Dead cells removal microbeads, Myltenyi) in presence of Citrate buffer. One Million cells per donor samples, healthy donor PBMCs and VeriCells were seeded in 96-well plate. Cells were washed, and each well was then stained with 100 μL of a unique double metal–labelled (Y89, Cd-106, Cd-110, Cd-112, Cd-116 and Cd-196) anti-CD45 antibody mix to further barcode the cells of individual donor. Cells were then washed twice, and five samples were combined into a single well. Cells were first stained with Fc block (BD Biosciences) during 10 minutes at room temperature. Cells were then washed and stained with the heavy metal–labelled antibody mixtures for 30 minutes on ice and 200 μM cisplatin during the last 5 minutes for the discrimination of live and dead cells. Cells were washed twice and fixed in 2% paraformaldehyde in PBS overnight at 4°C. Cells were then washed and resuspended in 250 nM iridium DNA intercalator (Fluidigm) in 2% paraformaldehyde/PBS at room temperature. Cells were washed, pooled together, and adjusted to 0.5 million cells per milliliter in MaxPar water together with 10 % equilibration beads (EQ Four element calibration beads, Fluidigm) for acquisition at 250 events/ second on a HELIOS mass cytometer (CyTOF, Fluidigm).

#### Data analysis

After mass cytometry acquisition, signals for each parameter were normalized based on EQ beads (Fluidigm). Each sample was manually de-barcoded followed by gating on live CXCR5^+^CD4^+^ T cells for T_FH_ analysis (CD45^+^ DNA^+^ cisplatin^−^ CD3^+^ cells) after gating out residual antigen-presenting cells (HLA-DR^+^CD3^-^ such as monocytes (CD14) and B cells (CD19) using FlowJo (Tree Star) software.

### Detection of SARS-CoV-2 specific memory B cells

Spike-specific B cells were detected using either sequential staining of biotinylated Recombinant SARS-CoV-2 Spike-Trimer (HEK) (Miltenyi) combined with streptavidin-PE or with probes already conjugated with Alexa Fluor 647 for Spike RBD (R&D Systems) and conjugated with Alexa Fluor 488 for Full-length spike protein (R&D Systems) (adapted from reference^24^). 2×10^6^ of cryo-preserved PBMC samples were transferred in a 96-well U-bottom plate. Cells were first stained with Fc block (BD Biosciences) for 15 min at room temperature. Cells were then washed and stained separately with either 100 ng of Spike Trimer alone or probe master mix containing 200 ng spike-A488, and 25 ng RBD-A647 for 1 hour at 4C. Following incubation with antigen probes, cells were washed twice and stained with Blue Live Dead (Thermofischer) during 10 minutes at room temperature. Cells were washed again and stained with anti-CD7, anti-CD14, anti-CD19, anti-CD20, anti-CD21, anti-CD24, anti-CD27, anti-CD38, anti-CD71, anti-IgD, anti-IgM, anti-IgG, anti-HLA-DR, and anti-CXCR5 for 30 minutes on ice. Cells stained with the Spike Trimer were fixed with the transcription factor buffer (Thermofischer) and intra-cellularly stained for IRF4 and Blimp-1. Cells stained with RBD and full Spike were fixed overnight in 1 % PFA. Samples were acquired on FACSymphony. All Antibodies were purchased from BD biosciences except towards Blimp-1 and IRF4 (Thermofischer) and IgM, CD71 and HLA-DR (Biolegend).

### Inflammatory markers

The following enzyme-linked immunosorbent assay (ELISA) kits were used according to manufacturer protocols. From R&D Systems: Human CD14 DuoSet ELISA (DY383), Human CD163 DuoSet ELISA (DY1607), Human LBP DuoSet ELISA (DY870-05), Human Galectin-9 DuoSet ELISA (DY2045), Human GDF-15 Quantikine ELISA Kit (DGD150), Human CXCL4/PF4 Quantikine ELISA (Kit DPF40), Human IFN-alpha (41100); from Ebioscience: Human MPO Instant ELISA Kit (BMS2038INST); from Thermo Scientific: Invitrogen novex IP 10 Human ELISA Kit (KAC2361);from Abcam Human C-Reactive Protein/CRP (Ab99995); from MyBioSource: Human zonulin ELISA Kit (MBS706368); from Meso Scale diagnostics: human Calprotectin (F21YB-3).

### Serology

A multiplexed bead-based flow cytometry assay, referred to as microsphere affinity proteomics (MAP), was adapted for detection of SARS-CoV-2 and the receptor-binding domain (RBD) antibodies as described^25-28^.

### Statistics

Comparative analyses of frequencies of cell subsets and marker expression between samples were performed using Wilcoxon rank sum tests, extended to Kruskal-Wallis tests by ranks for more than 2 levels in a grouping variable; resulting P values were adjusted for multiple testing using the Benjamini-Hochberg method to control the false discovery rate. Data are displayed as violin plots showing all data points (minimum to maximum). Correlations were calculated with the Pearson’s test. A correlation matrix was calculated comparing phenotypic and serological marker variables in a pairwise fashion, using the corr.test function from the psych CRAN package; the corrplot package was subsequently used to graphically display the correlation matrix. Resulting P values were adjusted for multiple testing using the Bonferroni method. Pearson’s correlation coefficients were indicated by a heat scale whereby blue color shows positive linear correlation, and red color shows negative linear correlation. All statistical analyses were performed using GraphPad Prism and R. Statistical significance was set at a threshold of *P < 0.05, **P < 0.01, ***P < 0.001, and ****P < 0.0001.

### Data and sample availability

The datasets generated and analysed during the current study are available from the corresponding author on reasonable request. The patient samples are not available on request due to restricting ethical and legal approvals.

## Extended data figures and tables

**Extended Data Fig. 1.**
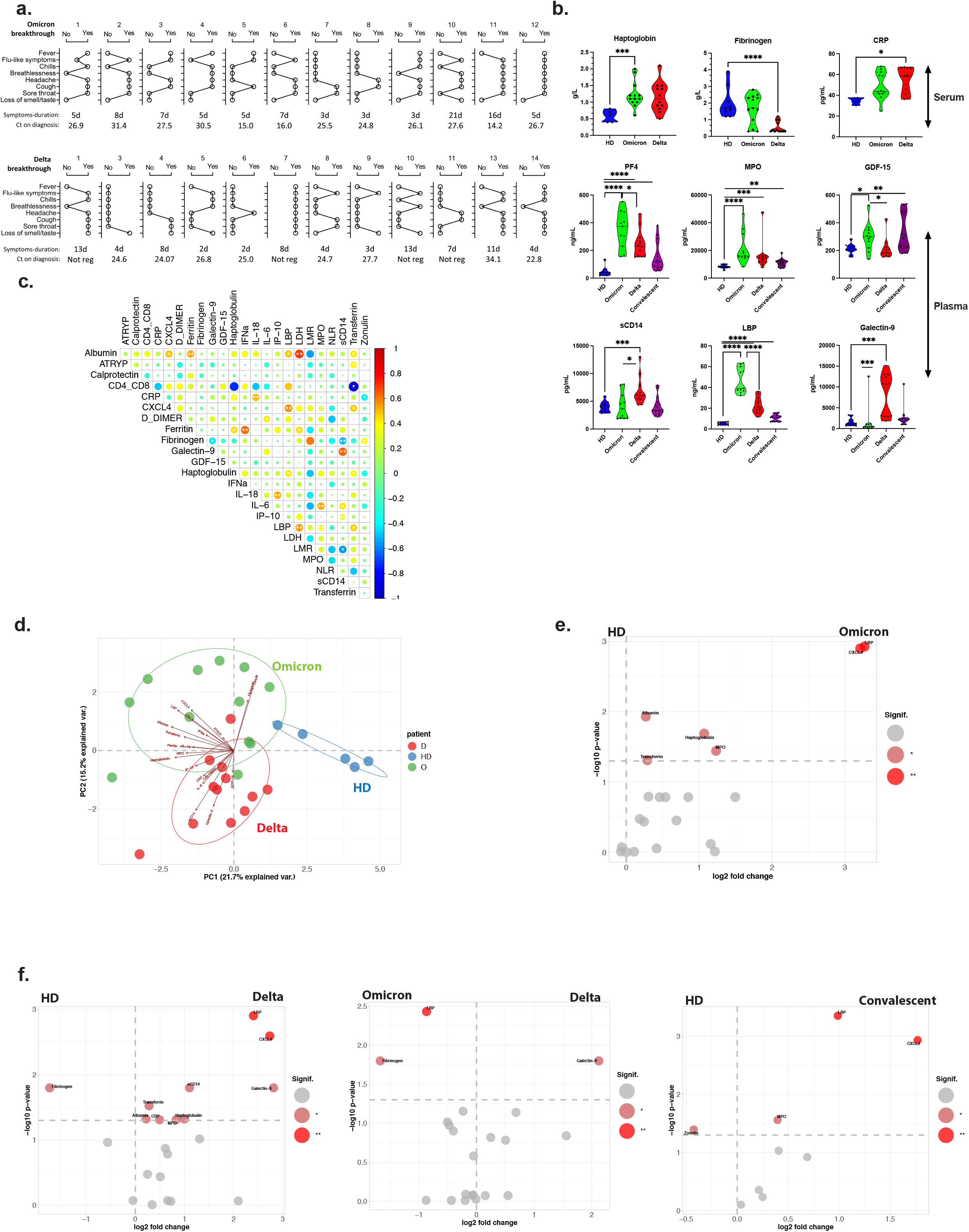
Symptoms and Inflammation during Omicron and Delta breakthrough infection. **a**. Self-reported symptoms of Omicron (top) and Delta (bottom) breakthrough COVID-19 cases at the time of inclusion. Duration of symptoms and PCR cycle threshold (Ct) values from the initial SARS-CoV-2 diagnostic tests are indicated. **b**. Acute phase proteins were measured in the serum (first row) and inflammatory markers were quantified in the plasma (second and third rows) of vaccinated individuals (either health care workers - healthy donors, 4 months since second dose of vaccine), Omicron breakthrough infected (8-13 d after symptom debut), Delta breakthrough infected (5-24 d), and unvaccinated convalescent COVID-19 patients (3-6 months post-infection). Statistical analyses were performed by Mann-Whitney Test and significant differences labelled with *, **, *** and **** for p<0.05, p<0.01, p<0.001 and p<0.0001 respectively. **c**. Systemic biological signature of included patients. Correlogram describes the potential interaction between the different molecules measured in the serum (anti-Trypsin, LDH, Albumin, Transferrin, Ferritin, Haptoglobin, CRP, IFNα, IL-6, IL-18), or in the plasma (Calprotectin, CXCL4, D-Dimer, Fibrinogen, Galectin-9, GDF-15, IP-10, LBP, MPO, sCD14 and Zonulin) and cellular ratio associated with COVID-19 in the peripheral blood such as CD4^+^/CD8^+^, LMR (Lymphocytes/ Monocytes Ratio), and NLR (Neutrophils/Lymphocytes Ratio). Cells counts were obtained by the staining of fresh blood in Trucount Tubes (BD Bioscience). The correlations are represented by a cold to hot heatmap (blue= negative correlation and red= positive correlation) and the significant combinations are indicated by an asterisk (* and ** for p<0.05 and p<0.01). **d**. Inflammatory signature of Omicron and Delta breakthrough infection. High dimensional analysis of individual patients according to the serological and plasma profile measured in **b**. Each circle represents one individual patient on the principal component analysis (PCA). Delta-and Omicron-infected cases are represented in red and green respectively and compared to vaccinated healthy donors (blue). **e**. Quantification of the Omicron inflammatory signature. Volcano plot summarized the significant fold change in the serum and plasma of Omicron cases for each marker and the significance of these fold changes. Statistically significant changes in marker expression is represented by different colours with * and ** for p<0.05 and p<0.01 respectively. **f**. Quantification of the Delta, Omicron and COVID-19 convalescent inflammatory signature. Volcano plot summarizes the significant fold change in the serum and plasma of Delta cases for each molecule and the significance of these fold changes in comparison with healthy donors (left) or Omicron-breakthrough cases (middle). COVID-19 convalescent patients were compared to vaccinated healthy donors (right). The statistically different molecules are represented in the graph with * and ** for p<0.05 and p<0.01 respectively.

**Extended Data Fig.2.**
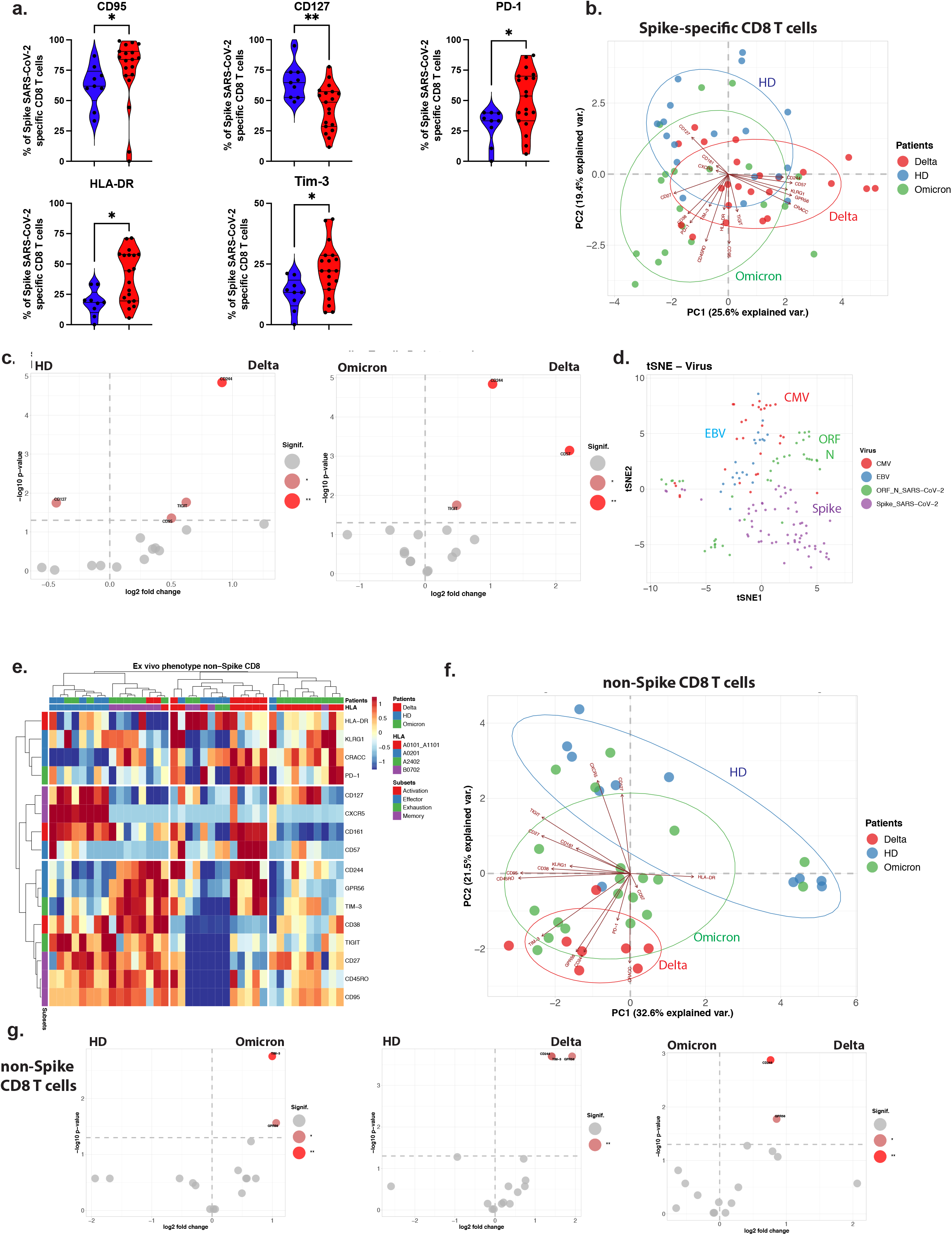
CD8+ T cell cellular immunity during Omicron and Delta breakthrough infection. **a**. Signature of Spike-specific CD8^+^ T cells during breakthrough Omicron infection. The expression of surface markers was compared between Spike-specific dextramers identified in healthy donors (blue) and in Omicron-infected cases (red). The statistically different molecules are represented in the graph with * and ** for p<0.05 and p<0.01 respectively (Mann-Whitney test). **b**. Signature of Spike-specific CD8^+^ T cells during breakthrough COVID-19 vs healthy donor controls. High dimensional analysis of individual patients according to the phenotype of Spike-specific Dextramers in principal component analysis (PCA) biplots. Each dot represents one individual patient analysis. Delta- and Omicron-infected patients are represented in red and green respectively and compared to vaccinated healthy donors (blue). **c**. Analysis of log P value vs fold change, volcano plots of Spike-specific CD8 T cells during Delta infection vs HD (left), and Delta vs Omicron (right). Significantly upregulated markers are indicated in darker shades of red. **d**. Distribution of SARS-CoV-2 specific T cells and bystander CD8^+^ T cells in tSNE plot. The phenotype of Spike-, Nucleocapsid-, and ORF-specific CD8^+^ T cells was compared to immune profile of CMV- and EBV-specific CD8^+^ T cells from our entire cohort. The profile of each virus-specific CD8^+^ T cell was visualized by tSNE representation with CMV-, EBV-, SARS-CoV-2 non-spike (ORF/N), and Spike-specific CTLs in red, blue, green and purple respectively. **e**. Immune phenotype *ex vivo* of non-spike (ORF/N) SARS-CoV-2 specific CD8^+^ T cells. A cold to hot heatmap represents the scaled frequency of each individual marker expressed by non-Spike antigen-specific CD8^+^ T cells. The distribution of markers and patients were automatically performed by unsupervised hierarchical clustering. Patient and HLA type are indicated in top two rows (Delta breakthrough COVID-19, Omicron breakthrough COVID-19, healthy donors (HD); HLA subtypes). The identification of marker types as subsets (Activation, Effector, Exhaustion, Memory) are indicated in the leftmost column. **f**. PCA Biplot analysis of non-Spike SARS-CoV-2 specific CD8^+^ T cells during breakthrough Delta and Omicron COVID-19 vs HD. **g**. Analysis of log p-value vs fold change, volcano plots of non-Spike SARS-CoV-2 specific CD8^+^ T cells during Omicron infection vs HD (left), Delta vs Omicron (middle) and Delta vs Omicron (right). Significant markers are indicated in darker shades of red.

**Extended Data Fig. 3.**
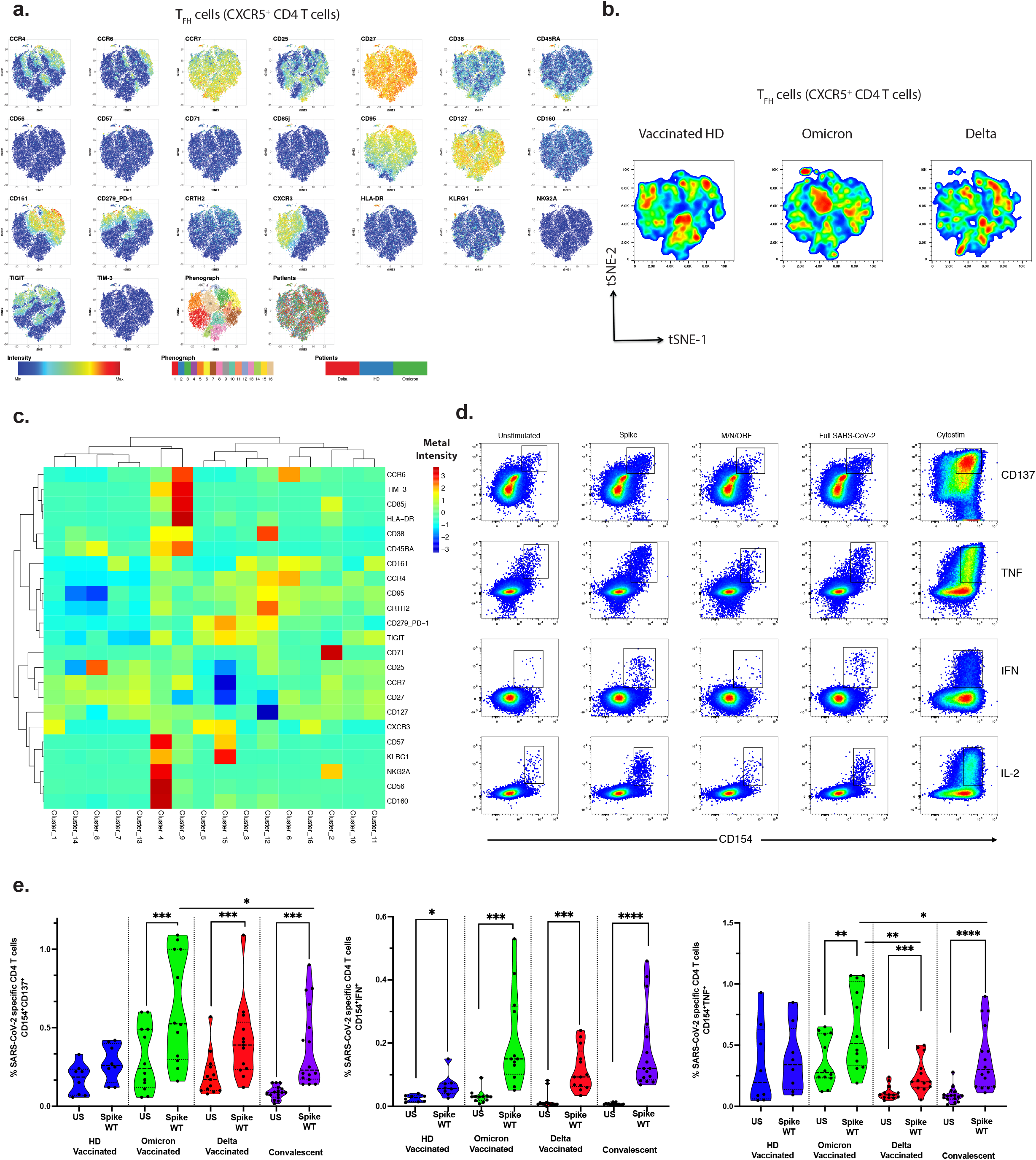
T helper cellular immunity during Omicron and Delta breakthrough infection. **a**. Phenotype of Follicular helper CD4 T cells during breakthrough infection. Mass cytometry staining of T_FH_ cells (identified as CXCR5^+^ CD4 T cells) in vaccinated infected patients. Cells from representative patients of each group (n=6) were concatenated and visualized by tSNE. The intensity of each marker is represented by a heatmap. The Phenograph algorithm was used to automatically identify the different clusters. Distribution of T_FH_ cells in tSNE plot is shown for markers as indicated. **b**. Overview and spatial visualization of T_FH_ cells from vaccinated non-infected, Omicron breakthrough infected and Delta breakthrough infected patients. Localization is based on specific co-expression of markers detailed in **c**. Immune phenotype *ex vivo* of T_FH_ cells. A cold to hot heatmap represents the scaled intensity of each individual metal-tagged marker expressed by T_FH_ cells. The distribution of markers and clusters were automatically performed by unsupervised hierarchical clustering. **d**. Identification of SARS-CoV-2 specific CD4 T cells by flow cytometry. Representative dot plots of CD4 T cells after stimulation with overlapping peptides coding for Spike, Membrane, Nucleocapsid and the entire SARS-CoV-2 proteom. Cytostim was used as positive control. Intra-cellular staining was performed after overnight stimulation and Live/Dead surface staining. CD4 T cells were gated as Live single cells lymphocytes co-expressing CD3 and CD4Quantification of SARS-CoV-2 specific CD4 T cells by flow cytometry. Cumulative frequencies of Spike SARS-CoV-2 specific CD4 T cells identified in vaccinated healthy donors, Omicron-or Delta-infected and in recovered COVID-19 patients. The statistical analysis was performed with Wilcoxon test on paired samples for the comparison between Unstimulated and peptides stimulated samples and with rank Mann-Whitney for the comparison between different groups. The statistically different molecules are represented in the graph with *, **, *** and **** for p<0.05, p<0.01, p<0.001 and p<0.0001 respectively.

**Extended Data Fig. 4:**
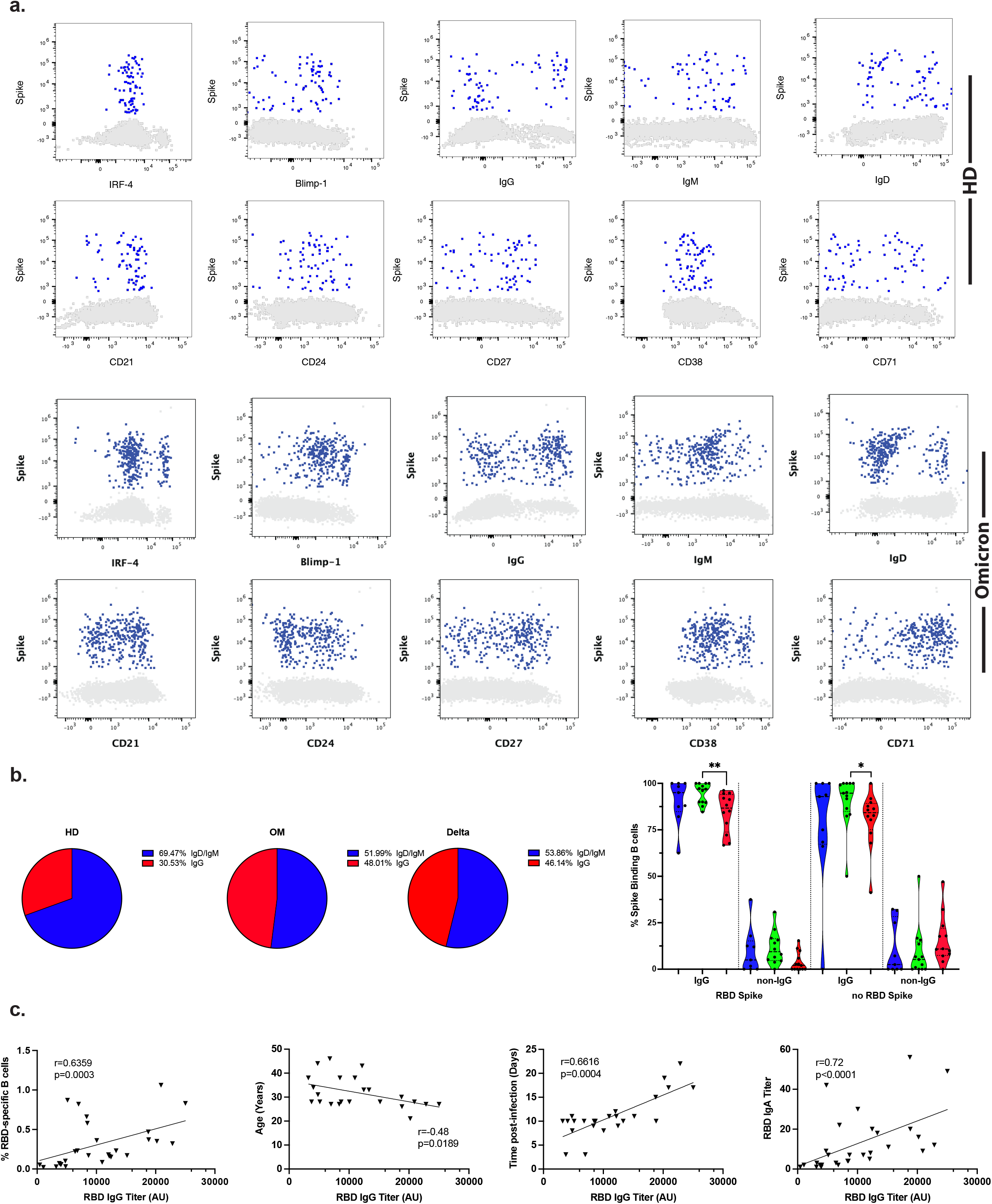
B cell and humoral immunity during Omicron and Delta breakthrough infection. **a**. Phenotype of Spike-binding B cells during breakthrough infection. Dot plots of live singlet total B cells from one representative healthy donor (top two rows) and one Omicron-infected individual (bottom two rows) are shown. Dark blue dots are Spike-binding B cells. Grey outlined plots show non-binding B cells. Surface molecules (IgG, IgD, CD21, CD24, CD27, CD38, CD71) and intra-cellular transcription factors (IRF-4 and Blimp-1) are represented on the X axis and Spike-binding on the Y axis. **b**. Distribution of class switched IgG vs IgM/D of Spike-binding B cells in pie charts as indicated. Spike binding B cells that are IgG vs Non-IgG are shown in violin plots. HD (blue), Omicron (green) and Delta (red) are shown for RBD-Spike binders (left) and non-RBD Spike binders (right). **c**. Humoral response and clinical parameters. The correlations between anti-RBD IgG titer and the frequency of Spike-binding B cells, the age of patients, the inclusion time post-infection and IgA titer were calculated by a Spearman’s rank order test, r values and P values are indicated on the graphs.

**Extended Data Fig. 5:**
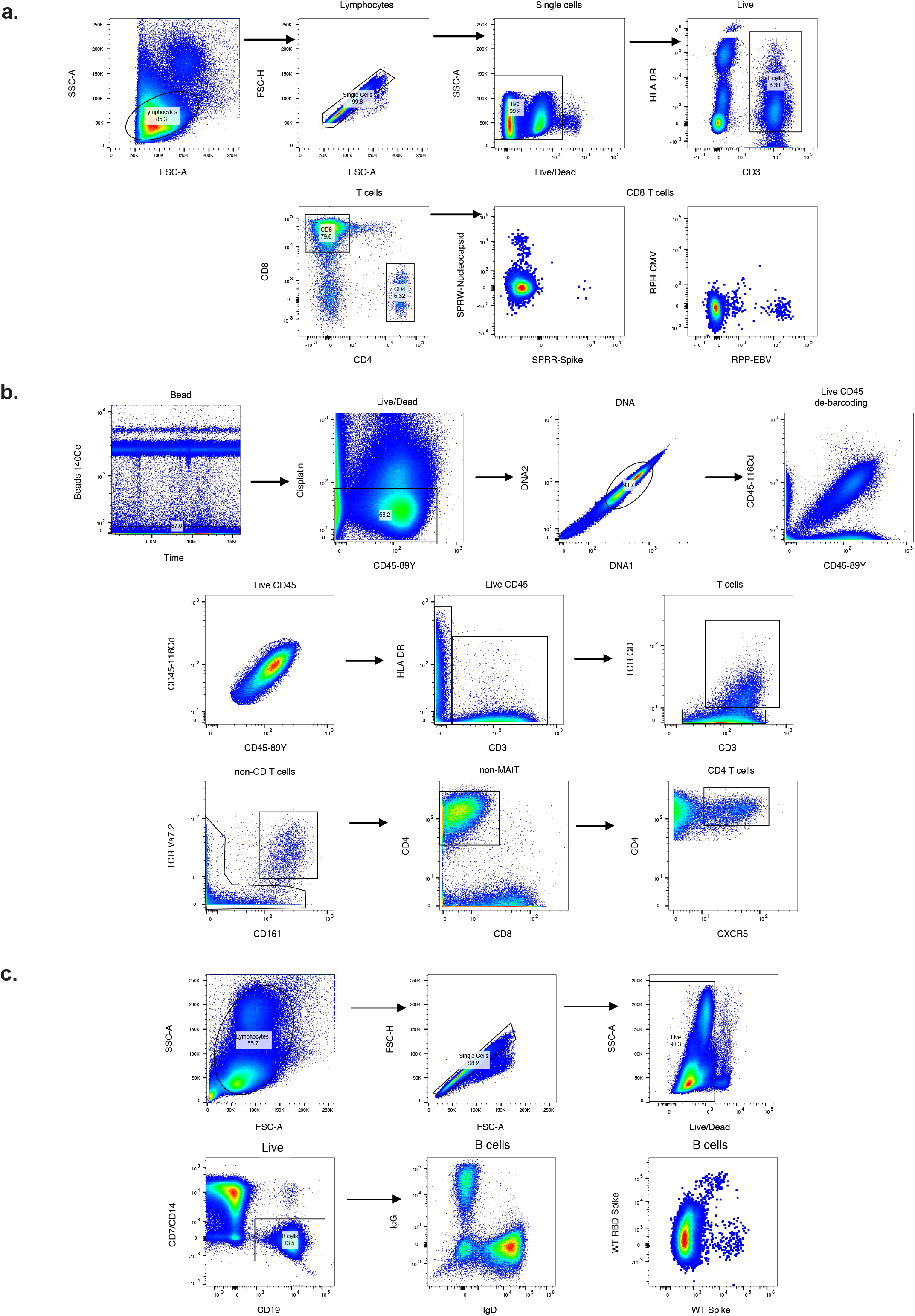
Gating strategy. **a**. Identification of Virus-specific CD8 T cells by flow cytometry. Illustrative example of Class I restricted multimers targeting Spike-, Nucleocapsid- and EBV-specific CD8 T cells is displayed. b. Identification of Follicular CD4 T cells by mass cytometry. Individual donor cells were de-barcoded according to their unique combination of double CD45 expression. TFH cells were identified in total CD4 T cells after exclusion of doublets, dead, non-T cells, γδ T cells and MAIT cells (CD161^+^TCR Vα7.2^+^). c. Identification of Spike-binding B cells by flow cytometry. Spike and Spike RBD SARS-CoV-2 binding B cells were identified in total B cells after exclusion of doublets, dead, and Lineage^+^ (Monocytes, NK cells and T cells). The phenotype of B cells was also studied by high parameters flow cytometry to detail naÏve (IgD^+^) and Switched (IgG^+^) memory B cells.

